# Heat inactivation of serum interferes with the immunoanalysis of antibodies to SARS-CoV-2

**DOI:** 10.1101/2020.03.12.20034231

**Authors:** Xiumei Hu, Taixue An, Bo Situ, Yuhai Hu, Zihao Ou, Qiang Li, Xiaojing He, Ye Zhang, Peifu Tian, Dehua Sun, Yongyu Rui, Qian Wang, Dan Ding, Lei Zheng

**Author notes:** Corresponding authors: Prof. Lei Zheng, or Prof. Dan Ding. These authors contributed equally to this work (Xiumei Hu, Taixue An, Bo Situ).

## Abstract

The detection of serum antibodies to the severe acute respiratory syndrome coronavirus 2 (SARS-CoV-2) is emerging as a new tool for the coronavirus disease-2019 (COVID-19) diagnosis. Since many coronaviruses are sensitive to heat, heating inactivation of samples at 56 °C prior to testing is considered a possible method to reduce the risk of transmission, but the effect of heating on the measurement of SARS-CoV-2 antibodies is still unclear. By comparing the levels of SARS-CoV-2 antibodies before and after heat inactivation of serum at 56 °C for 30 minutes using a quantitative fluorescence immunochromatographic assay, we shown that heat inactivation significantly interferes with the levels of antibodies to SARS-CoV-2. The IgM levels of all the 34 serum samples (100%) from COVID-19 patients decreased by an average level of 53.56%. The IgG levels were decreased in 22 of 34 samples (64.71%) by an average level of 49.54%. Similar changes can also be observed in the non-COVID-19 diseases group (n=9). Of note, 44.12% of the detected IgM levels were dropped below the cut-off value after heating, suggesting heat inactivation can lead to false-negative results of these samples. Our results indicate that heat inactivation of serum at 56 °C for 30 minutes interferes with the immunoanalysis of antibodies to SARS-CoV-2. Heat inactivation prior to immunoanalysis is not recommended and the possibility of false-negative results should be considered if the sample was pre-inactivated by heating.

## Introduction

The current outbreak of coronavirus disease-2019 (COVID-19) caused by a novel severe acute respiratory syndrome coronavirus 2 (SARS-CoV-2) is posing a serious threat to public health.^1-3^ Early diagnosis of suspect cases is critical to reduce and interrupt the transmission of COVID-19 from person-to-person.^4^ Currently, laboratory testing of viral nucleic acid by real-time reverse transcriptase–polymerase chain reaction (RT-PCR) assay is the “gold standard” for COVID-19 diagnosing.^5^ However, the requirement of sophisticated instruments and laboratory conditions, tedious experimental procedures, and longer detection time significantly hamper its widespread applicability.^4^ Antibodies produced in the blood after COVID-19 infection are emerging as a promising class of biomarkers.^6^ The antibodies to SARS-CoV-2 are specific, sensitive, and more importantly, their detection can be much faster and simpler than RT-PCR, which allows rapid screening of suspect cases to be possible.^7^

All the biological specimens for COVID-19 testing should be considered to be potentially infectious. Therefore, test must be performed by medical professionals with protective equipment in a qualified laboratory. To further reduce the risk of exposure to infectious agents, viral inactivation before sample handling are usually be recommended.^8,9^ While the sensitivity of SARS-CoV-2 to the conditions of inactivation is unknown, it is reported that many coronavirus such as SARS are heat-sensitive and can be killed at 56 °C for 30 minutes.^10-14^ It is thus inferred that heating at 56 °C could be an effective approach for SARS-CoV-2 inactivation.^15^ However, the effect of heating at 56 °C on COVID-19 antibody detection is unclear. The objective of this study was to compare the levels of COVID-19 antibody before and after heat inactivation.

## Methods

A total of 34 serum samples with positive SARS-CoV-2 antibody results from patients with COVID-19 infections and 9 serum form non-COVID-19 diseases were collected form Hankou Hospital, Wuhan city with approval of the ethics committee (hkyy2020-004). All patients with COVID-19 infections were confirmed by RT-PCR. The antibody detection kits for SARS-CoV-2 were obtained from KingFocus Biomedical engineering Co.,Ltd (AIE/Quantum dot-based fluorescence immunochromatographic assay, AFIA). The immunoassay quantitatively measures IgM and IgG antibodies to SARS-CoV-2. Serum samples before and after heat inactivation at 56 °C for 30 minutes were analyzed according to the protocol. Briefly, 100 microliters of serum was dropped on the test card and the fluorescence signal was measured after 15 minutes. Detection values above the cut-off threshold are considered positive.

## Results

In the patients with COVID-19, the IgM signals of all the 34 serum samples (100%) decreased (**Figure 1**) by an average level of 53.56% ([95% CI, 7.64%-99.49%]; *P*□< □0.013) after heat inactivation. The IgG signals were decreased in 22 of 34 samples (64.71%) by an average level of 49.54% ([95% CI,8.76%-90.32%]), and 12 samples (35.29%) increased with a median percentage of 24.22%. 44.12% of the IgM signals form COVID-19 patients were below the cut-off value after heat inactivation. In the non-COVID-19 group, the IgM levels were decreased in 7 of 9 samples (77.78%) by an average of 43.31% (**Figure 2**) after heat inactivation and 2 samples (22.22%) increased with an average level of 29.84% ([95% CI,5.44%-54.23%]). The IgG signals were decreased in 7 of 9 samples (77.78%) by an average level of 79.42% ([95% CI,44.54%-114.31%]), and 2 samples (22.22%) increased with an average level of 44.00% ([95% CI,21.37%-66.63%]). None of the measured antibodies became higher than the cut-off value after heating.

**Figure 1.**
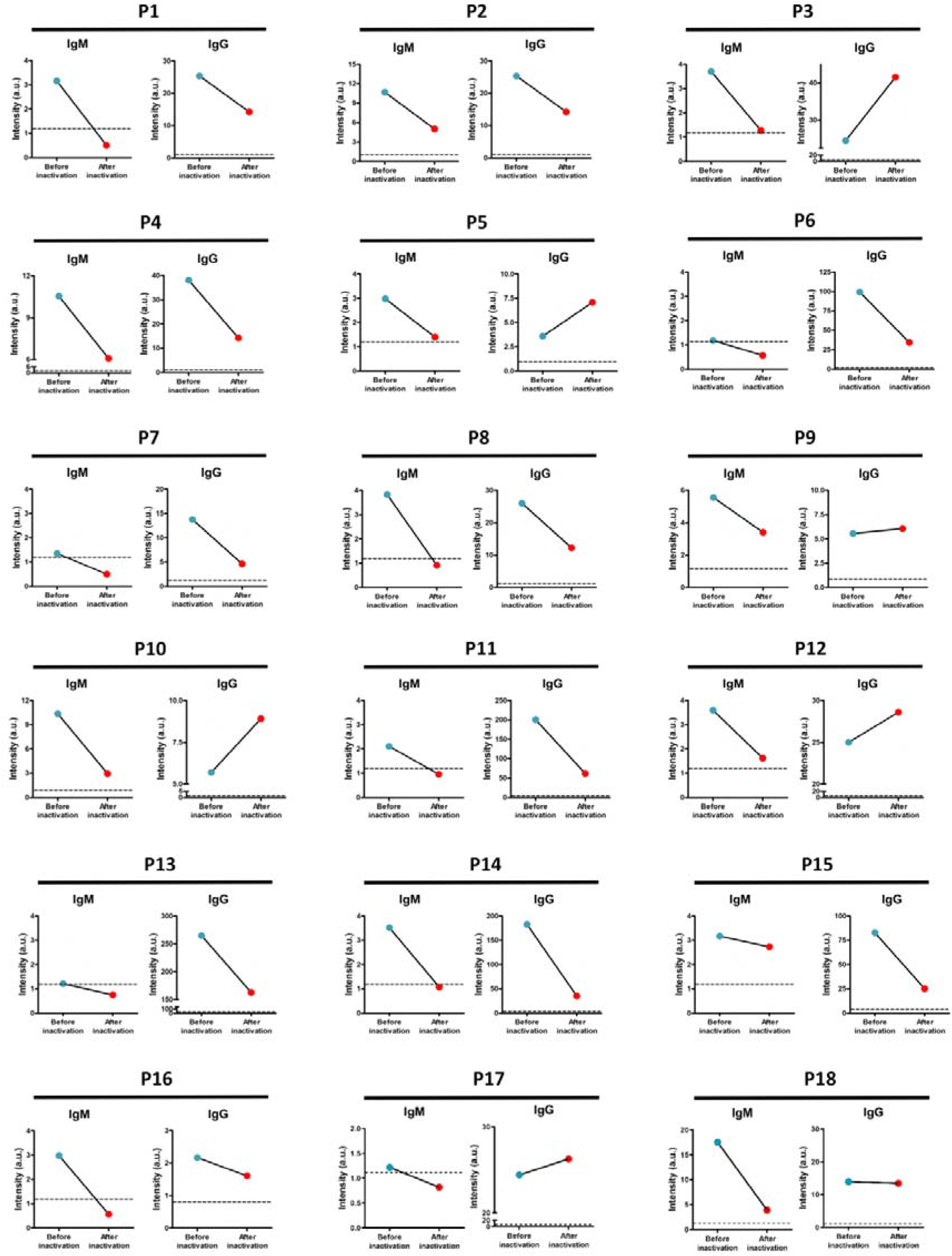
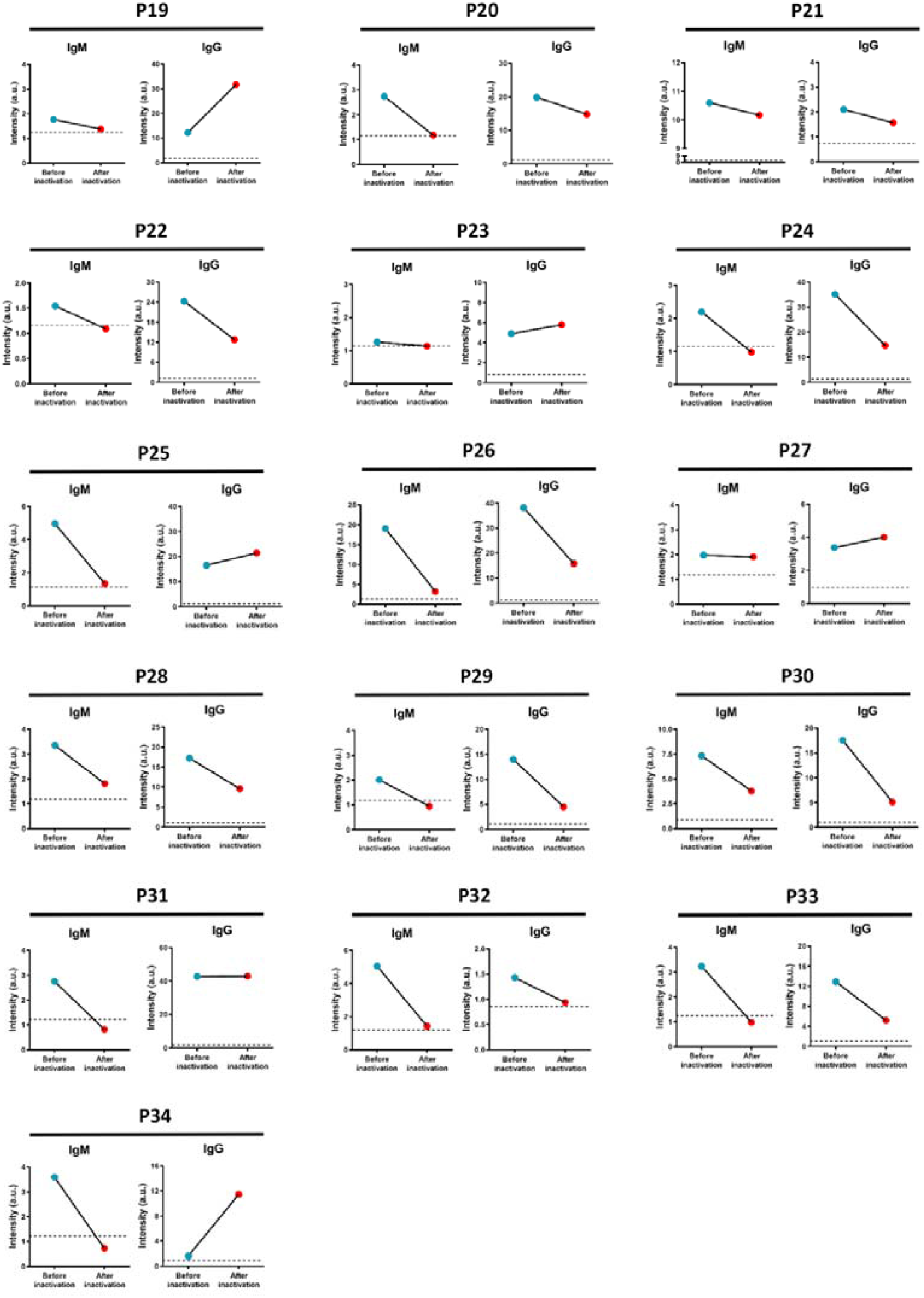
The changes in the IgM and IgG levels of the 34 serum samples from patients with COVID-19 infections detected by AFIA before and after heat inactivation. Dash line indicates the cut-off value of the assay.

**Figure 2.**
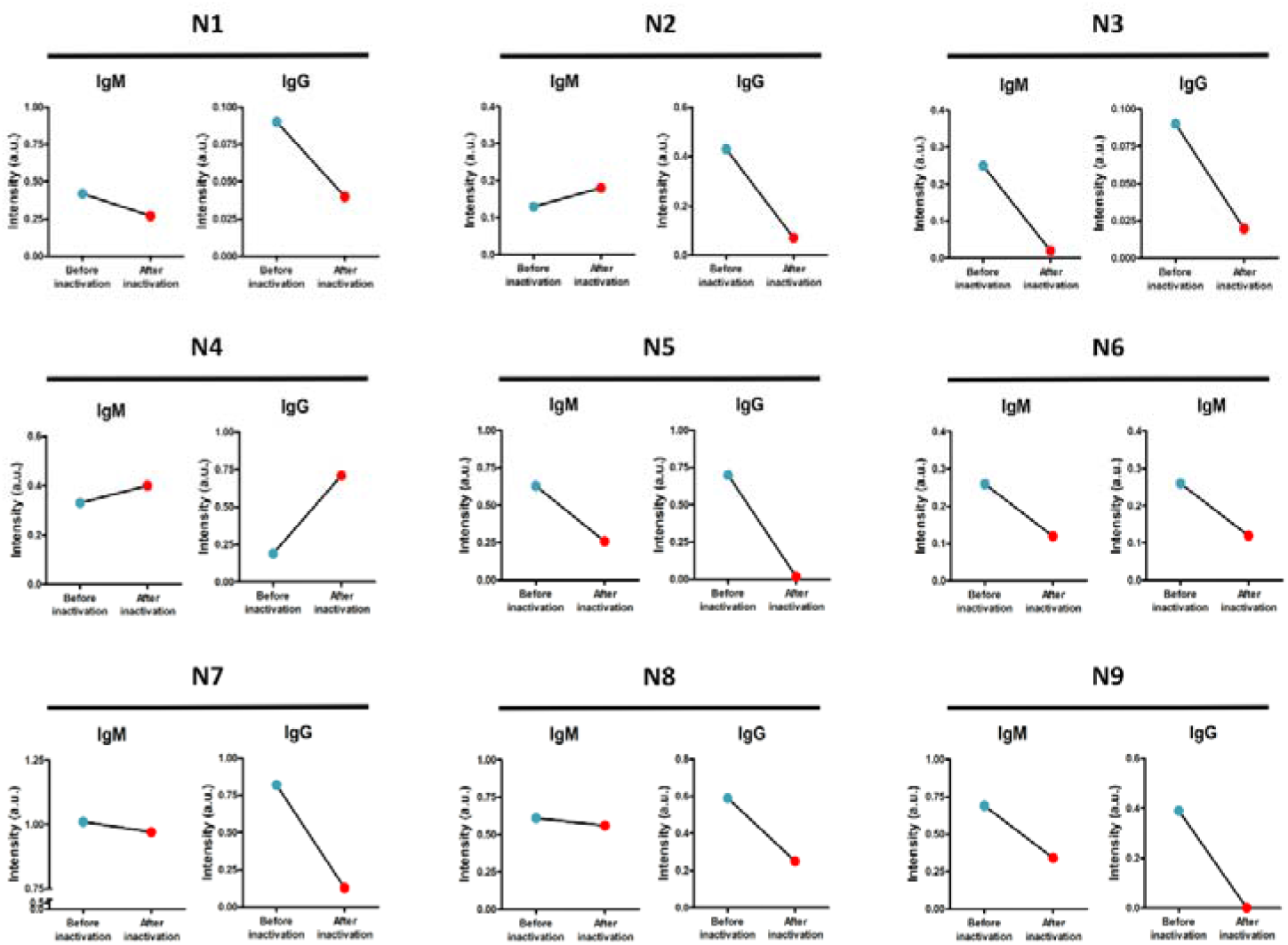
The changes in the IgM and IgG levels of the 9 serum samples from non-COVID-19 group detected by AFIA before and after heat inactivation.

## Discussion

This study analyzes the changes in SARS-CoV-2 antibody concentration before and after heat inactivation at 56 °C for 30 minutes. We found that after heat inactivation, all of the serum IgM (100%) demonstrated significantly lower levels. For IgG, 64.71% of the sample levels dropped after heat inactivation.

All immunological assays are highly dependent on the recognition and binding of antigens to antibodies. The decrease in SARS-CoV-2 antibody levels may be related to their structural change of denaturation and aggregation.^16,17^ Previous studies have shown that antibodies can be denatured and lose their antigen binding activities after heating,^18^ and IgM is reported to be less thermally stable than IgG^19,20^ due to their different compositions and structures of heavy chains.^21^ This is consistent with our results that SARS-CoV-2 IgM concentration decreased more significantly than IgG after heating. In addition, the IgG levels in 12 samples (35.29%) increased with a median of 24.22% after heating, which may be due to the increases immunogenicity caused by the formation of IgG aggregates heating at 56 □.^16,22,23^ It is noteworthy that after heat inactivation, 44.12% of the IgM levels form COVID-19 patients were below the cut-off value. These results suggests that heat inactivation of serum can lead to false-negative results in these samples.

## Conclusions

Heat inactivation of serum at 56 °C for 30 minutes interferes with the immunoanalysis of antibodies to SARS-CoV-2. For highly suspected cases, the possibility of false-negative results should be considered if the sample was inactivated by heating.

## Data Availability

The data used to support the findings of this study are available from the corresponding author upon request.

